# Development and validation of a methodology to measure exhaled carbon dioxide (CO_2_) and control indoor air renewal

**DOI:** 10.1101/2022.09.22.22280262

**Authors:** Marta Baselga, Juan J. Alba, Alberto J. Schuhmacher

## Abstract

The measurement of CO_2_ has positioned itself as a low-cost and straightforward technique to indirectly control indoor air quality, allowing the reduction of the concentration of potentially pathogen-loaded aerosols to which we are exposed. However, on numerous occasions, bad practice limits the technique for CO_2_ level interpreting and does not apply methodologies that guarantee air renewal. This work proposes a new methodology for measuring and controlling CO_2_ levels for indoor air in shared spaces. The proposed methodology is based on three stages: diagnosis, correction protocols, and monitoring/control/surveillance (MCS). The procedure is explained using a cultural center as an actual base case study. Additionally, the procedure was validated by implementing 40 voluntary commercial spaces in Zaragoza (Spain). Standardization of methods is suggested so that the measurement of CO_2_ becomes an effective strategy to control the airborne transmission of pathogens and thus prevent future Covid-19 outbreaks and novel pandemics.

## 1. Introduction

Since the global acceptance of the transmission of COVID-19 by aerosols,^1^ numerous strategies have been developed to prevent air quality degradation by different means. It is possible to differentiate between air renewal and air purification strategies. On the one hand, renewal techniques evaluate and optimize air renewal in closed spaces. On the other hand, purification methods are varied and struggle to eliminate potential airborne pathogens. Although both strategies share the common goal of reducing the airborne spreading of respiratory diseases, air purification is seriously handicapped because it usually requires higher economic investments (e.g., bipolar ionization systems, UV-C radiation devices), and its efficacy has been poorly tested against SARS-CoV-2. As an exception, high-efficiency portable air cleaners (e.g., HEPA filters) are presented as an affordable, high-performance method that can filter up to 99.9% of submicron particles and reduce SARS-CoV-2 aerial viral viral viral load up to 80%.^2,3^ However, in actual operating conditions, it is necessary to consider the workflow of the equipment and its location to take advantage of its performance.

Ideally, measuring aerosols would allow knowing the quality of the air and estimating the risk of contagion. Nevertheless, the direct measurement of aerosols is highly complex and expensive since it requires specialized equipment. To overcome these hurdles, the carbon dioxide (CO_2_) measurement has been suggested as an indirect indicator of the risk of respiratory infectious diseases transmission.^4^ The measure of exhaled CO_2_ was proposed before the pandemic to control the airborne spread of diseases since it represents a greater probability of breathing air previously inhaled by other people.^5^ Consequently, the indoor CO_2_ measurement is suggested as a reasonable ventilation proxy. It is possible to determine what percentage of the exhaled air by another individual (*y*) according to the expression *y*= *Ce x* + *Ca*(1 −*x*). Where, *Ce* corresponds to the concentration of CO_2_ in exhaled air (estimated at 40,000 ppm), *Ca* to the ambient CO_2_ concentration and *x* to the fraction of exhaled air. For instance, assuming a basal value of 440 ppm (fresh air outdoors), which increases indoors up to 2,000 ppm, the approximate percentage of air that those individuals have already breathed will be 3.9 %.

One of the significant limitations of CO_2_ level measurement is that it cannot be directly related to the concentration of aerosols in the environment since the generation of bioaerosols depends on the type of respiratory activity.^6^ For example, aerosols generated during sustained vocalization (e.g., singing) are not comparable to those generated during silent breathing.^7–10^ Nevertheless, CO_2_ concentration limits have been proposed to reduce COVID-19 transmission, usually between 700 and 1,000 ppm, regardless of the event.^11,12^

SARS-CoV-2 is mainly spread over time and distance. An average of 3.1 ± 2.9 copies/L of air viral load average is deduced from a total of 313 samples.^13–35^ It has been established that close contact transmission is predominant, while fomites and aerosols explain special propagation events in punctual events, according to CDC guidelines.^36^ The massive infections associated with superspreading represent a scenario of complex epidemiological management. To date, superspreading events have only been reported indoors.^28,37–55^ Due to the difficulty of discriminating between superspreaders and non-spreaders,^28,56^ it is imperative to improve air quality in public and shared spaces. By implementing the proper measures, it is possible to reduce cases of respiratory diseases in the community and prevent future pandemics with devastating effects.

Many countries have implemented CO_2_ measurement as a standard in different spaces, such as shopping centers,^57^ collective transport,^58–61^ workspaces,^62^ or university and school classrooms.^62–67^ Beyond research projects, some governments have decided to bet on scientific criteria and invest resources to mitigate contagion among the population. For example, the U.K. government distributed more than 300,000 CO_2_ meters in its schools to optimize air renewal during classes.^68^ In Germany, a global approach was also carried out, investing more than €17M to guarantee the presence of meters in schools.^69^ An investment of $350 billion for state and local governments has recently been reported and $122 billion for schools, which can support making ventilation and air filtration upgrades in the U.S. to reduce the respiratory pathogens spreading.^70^

However, there are still no standardized methodologies for measuring and controlling CO_2_. “Aireamos” (in Spanish, it refers to “renewing air”),^71,72^ as an international independent scientific working group, has been a pioneer in developing the bases for measurement, making available to the population a set of valuable tools and resources to control the renewal of indoor air. However, there is still no straightforward procedure for intervening in public and shared spaces. Before the pandemic, CO_2_ measurement was mainly explored in school settings. Numerous investigations pointed to an improvement in the pupil’s concentration in well-ventilated classrooms.^73–75^

This article proposes a new methodology for effectively measuring and controlling CO_2_ in closed and shared spaces. The methodology is based on three stages: diagnosis, correction, and controlled monitoring. The methodology allows us to interpret the air renewal patterns and estimate if the renewal is appropriate to the activity and type of space. Additionally, this method has been validated as a pilot project in a 3-floors building with multiple teaching rooms and cultural activities and a subset of 40 local businesses in Zaragoza (Spain).

## 2. Materials and Methods

### 2.1. General description of the methodology

A methodology based on three standard stages with specific goals was developed. They enumerate as follows:

a. Diagnosis. It is intended to acquire detailed knowledge on how CO_2_ concentration levels vary within the physical limits of the shared space and what is its time evolution as a function of specific activities carried out in the shared space. The initial diagnosis includes a description of the space indicating the measures of the shared space (e.g., size, volume, and architectural geometry), other CO_2_ sources (e.g., gas stove), the presence and ubication of windows and doors, as well as the availability of Heating, Ventilation, and Air Conditioning (HVAC) systems and hoods. During this process, it is necessary to identify the “person in charge” and perform minimal instruction about the project and concepts such as air renewal, ventilation, metabolic CO_2,_ and how to measure it.
b. Correction. According to the acquired knowledge in the diagnosis stage, it is possible to proceed with the design of customized protocols and procedures to improve the air renovation that will imply variations in the CO_2_ concentration. The CO_2_ levels will be monitored with a CO_2_ meter to avoid trespassing a defined threshold considered as a low-risk level by the scientific community.
c. Monitoring, control, and surveillance (MCS): Continuous monitoring of CO_2_ concentration is performed to guarantee that the shared space is always kept at lowrisk levels. In case of the CO_2_ concentration levels reach the maximum level, an alert is issued so that people in charge of the shared space activate the protocols and procedures defined in the previous stage to fix the situation.

For each stage, achieving the primary goals depends on specific characteristics of the shared space, such as main business or social activity, geometrical properties, number and location of windows and doors, or a maximum number of people sharing the room at the same time. A case study and a practical application is presented in this article to explain this methodology.

### 2.2. Measurement of metabolic CO_2_

Any time measuring metabolic CO_2_ levels is required, Aranet CO_2_ meters (Aranet Wireless Solutions España SL, Spain) and Signos CO_2_ meters (Signos.io, Spain) will be used. The technical characteristics of these measuring devices are shown in Table 1.

**Table 1.** Technical characteristics of Aranet and Signos SIO_2_ CO_2_ meters.

| Characteristics | Aranet 4 Pro | Signos (SiO<sub>2</sub>) |
| --- | --- | --- |
| CO<sub>2</sub> measurement | < 9,999 ppm | < 5,000 ppm |
| Temperature measurement | 0 – 50 °C | -60 – 80 °C |
| Humidity measurement | 0 – 85 % | 0 – 95 % |
| Atmospheric pressure | 0.3 – 1.1 atm | 0.3 – 1.1 atm |
| Sensor | N-DIR | N-DIR |
| Sampling frequency | 1 min | 11 min |
| Transmission | Bluetooth (-12–4 dBm) | LoRa |
| Precision | ± 50 ppm | ± 50 ppm |
| Dimensions / weight | 70x70x24 mm / 104 g | 100x100x23 mm / 170 g |

### 2.3. The methodology explained in a case study

As a case study, the methodology for improving air quality was applied in a cultural center. The building has 1,200 m^2^ divided in three floors (Figure 1a). A 132 m^2^ room located in the basement was selected (Classroom 1, see Figure 1b).

**Figure 1.**
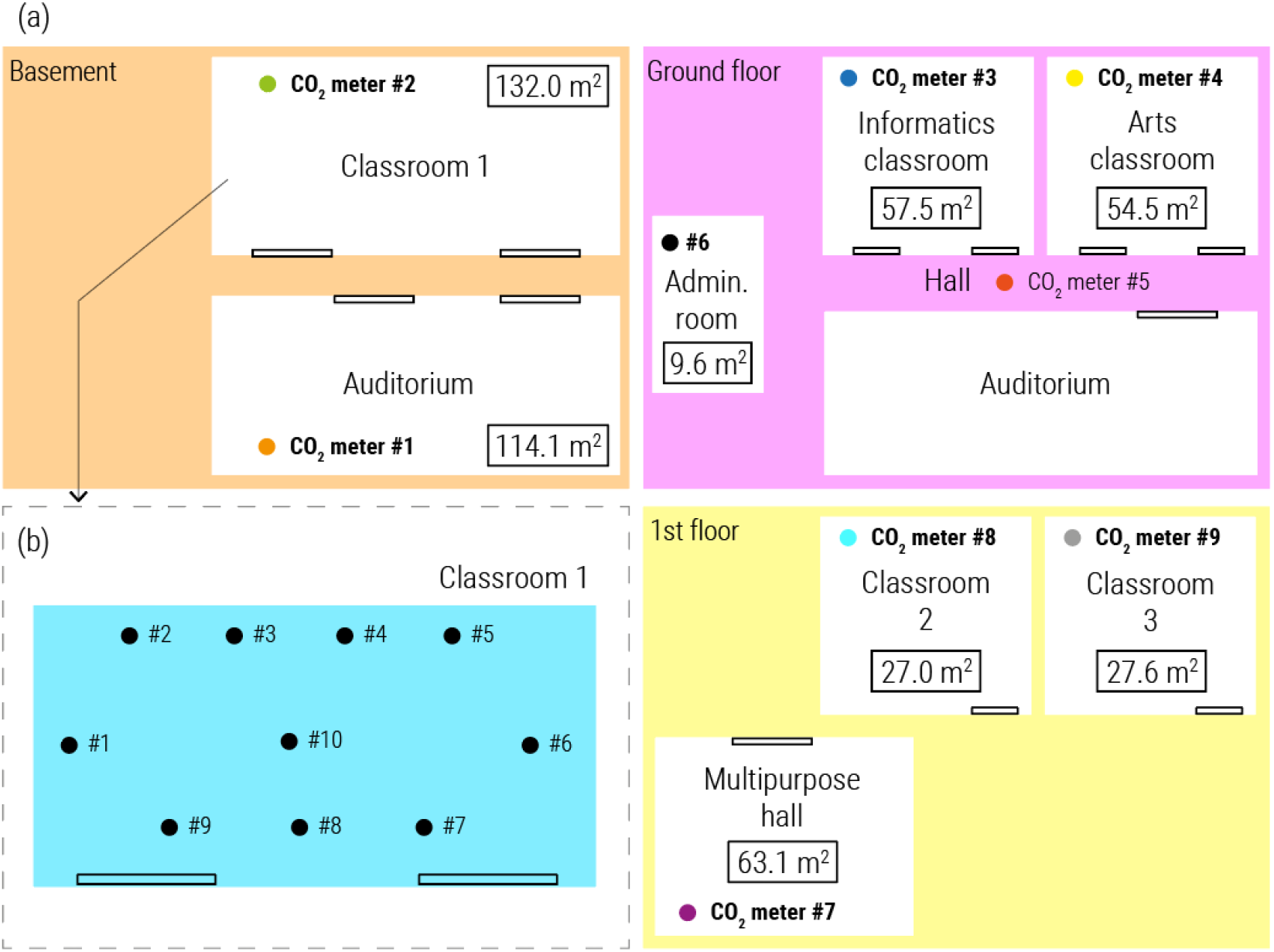
(a) General distribution of CO_2_ meters in the education building and (b) classroom 1 detail.

a. Diagnosis: The shared spaces were analyzed for four days (February 24^th^ to 27^th^, 2021) by placing 10 CO_2_ meters (Aranet 4) in strategic locations, as shown in Figure 1a. A CO_2_ measuring device was placed outdoors so that it was possible to calculate the increase in CO_2_ (Δ*Co*_2_) associated with human activity according to Equation 1; where (CO_2_)_ext_ corresponds to CO_2_ outside and (CO_2_)_ind_ to CO_2_ indoors.

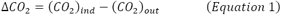

In the first approach, the data showed frequent high CO_2_ concentration levels probably linked to specific activities. Subsequently, the shared spaces were carefully monitored for six more days (February 27^th^ to March 5^th^, 2021) to confirm the hypotheses that could be made after the initial analysis. This process makes it possible to characterize the air renewal patterns in each space based on activity and capacity. At least one CO_2_ meter was placed every 20 m^2^ at a minimum height of 1.5 m. Ideally, they will be placed near exhaust fans (to measure a significant air sample) and/or in locations where air turnover is poor (e.g. corners), and far from windows and doors.
b. Correction: Based on the obtained results, an attempt was made to reduce the level of metabolic CO_2_ below 800 ppm. One of the classrooms in the basement of the building (Classroom 1) showed inefficient ventilation patterns. Dry ice was used to saturate the room with CO_2_ and find effective strategies to improve air renewal. According to Figure 1b, 10 CO_2_ meters (Aranet 4) were placed in the classroom to analyze the improvement of changes in the HVAC and natural ventilation (through doors and windows opening or closing).
c. MCS: This stage was performed until May 15^th^, 2022 using 10 CO_2_ meters (Signos). Whenever the cultural center exceeds the 800 ppm level for more than 30 minutes, it receives a telematic alert or phone call to ask the “person in charge” to correct the situation.

### 2.4. Broader validation in commercial spaces

Subsequently, the method was applied to 40 businesses / retail stores spaces located in Zaragoza’s city center (Spain). These included: one optician, twelve clothing stores, two kids’ clothing stores, five shoe stores, two jewelry stores, four food stores, two restaurants, five home stores, two consultancies, two dietary and health centers, and two florists. The selection of commercial spaces was made to cover different typologies. For example, customers spend long periods in restaurants, while their exposure to indoor air in flower retailers is minor. Two CO_2_ meters were placed in each establishment, except for a kid’s clothing store, where only one was placed, and another children’s clothing store, where three were placed.

Interventions in shared spaces were carried out following the three phases: diagnosis, correction, and MCS.

a. Diagnostic. Two CO_2_ meters (Aranet 4) devices were placed at strategic points for 2 or 3 days in each business. These meters were chosen because they record the CO_2_ level every minute. The CO_2_ curves were analyzed during the business hours of each establishment. This phase allowed us to know if the sensors took representative measurements of the total space and have a global image of the air quality in the volume area. The diagnostic process might include the use of risk calculators or methods for simulating disease propagation via aerosols based on the standard Wells-Riley model for infectious respiratory disease transmission by airborne particles.^71,72,76^
b. Correction. In spaces where it was necessary (typically presenting CO_2_ levels >1,000 ppm for more than 30 minutes), a correction phase was carried out where preventive measures were implemented to improve indoor air renewal. In most cases, this phase was superimposed on monitoring to determine the advantage obtained at the time.
c. MCS. The monitoring stage occurred during January and February 2022. For this stage, CO2 (Signos) meters were installed in each establishment. These CO_2_ meters, include IoT technology, enabled remote access to streaming data.

## 3. Results

### 3.1. The diagnostic stage to allow detection of inefficient air renewals

The diagnosis must be performed on a long enough time interval to cover the specific and regular casuistry of the establishment. For example, in the base case study (cultural center), the classrooms usually are occupied for daily classes. Therefore, a selection of days where the classrooms are occupied is sufficient. In this case, four days were used for the preliminary analysis of the space. Assuming a proper maximum CO_2_ level of 800 ppm, a maximum increase of 400-450 ppm compared to fresh outdoor air was considered acceptable.

As shown in Figure 2, there are peaks in the increment of CO_2_ associated with the activity of each room. Knowing the activity and the number of occupants is essential to interpret the CO_2_ measurements. Determining the ppm/person ratio can be helpful, as we proposed in previous work (see ^61^). This value incorporates the occupancy variable and allows a more precise definition of the efficiency of air renewal, although it assumes homogeneity in exhalation between individuals. In addition, it is crucial to request the usual activity in the space. Those responsible must not consciously increase ventilation (e.g., opening windows) to avoid underestimating CO_2_ measurements. A typical table with essential information is shown in Table 2. The method to fill in the table must be explained in the training phase since it is of utmost importance to interpret the data properly later. In the base case, the room had no open windows, except for the Hall and the Art Classroom. In contrast, all doors were open during diagnostic measurements.

**Table 2.** Example of a data sheet for CO_2_ measurements interpretation.

| Day | Duration | Activity | Occupation | Location | Mean<br>$\Delta\text{CO}_2$ | Max<br>$\Delta\text{CO}_2$ | Mean<br>ppm/person | Max<br>ppm/per-<br>son |
| --- | --- | --- | --- | --- | --- | --- | --- | --- |
| 1<sup>st</sup> day | 1:30 h | English class | 8 pax | Auditorium | 106 ppm | 196 ppm | 13.3 ppm | 24.5 ppm |
| 2<sup>nd</sup> day | 2:00 h | History class | 15 pax | Auditorium | 164 ppm | 234 ppm | 11.0 ppm | 15.6 ppm |
| 3<sup>rd</sup> day | 2:00 h | English class | 9 pax | Auditorium | 116 ppm | 170 ppm | 12.9 ppm | 18.9 ppm |
| 1<sup>st</sup> day | 1:30 h | English class | 10 pax | Classroom 1 | 89 ppm | 153 ppm | 8.9 ppm | 15.3 ppm |
| 3<sup>rd</sup> day | 2:00 h | English class | 8 pax | Classroom 1 | 76 ppm | 140 ppm | 9.5 ppm | 17.5 ppm |
| 4<sup>th</sup> day | 2:30 h | English exam | 18 pax | Classroom 1 | 219 ppm | 312 ppm | 12.2 ppm | 17.3 ppm |
| 4<sup>th</sup> day | 4:00 h | English exam | 7 pax | Informatics | 328 ppm | 751 ppm | 46.9 ppm | 107.3 ppm |
| 2<sup>nd</sup> day | 3:00 h | Art class | 6 pax | Art classroom | 68 ppm | 121 ppm | 11.3 ppm | 20.2 ppm |
| 3<sup>rd</sup> day | 1:30 h | Art class | 6 pax | Art classroom | 34 ppm | 125 ppm | 5.8 ppm | 20.8 ppm |
| 3<sup>rd</sup> day | 3:00 h | Art class | 6 pax | Art classroom | 79 ppm | 243 ppm | 13.2 ppm | 40.5 ppm |
| 3<sup>rd</sup> day | 2:30 h | Art class | 4 pax | Art classroom | 58 ppm | 118 ppm | 9.7 ppm | 19.7 ppm |
| 1<sup>st</sup> day | 2:30 h | English class | 7 pax | Multi. hall | 163 ppm | 263 ppm | 23.3 ppm | 37.6 ppm |
| 2<sup>nd</sup> day | 1:00 h | English class | 10 pax | Multi. hall | 181 ppm | 213 ppm | 18.1 ppm | 21.3 ppm |
| 3<sup>rd</sup> day | 2:30 h | English class | 8 pax | Multi. hall | 177 ppm | 279 ppm | 22.2 ppm | 34.9 ppm |

**Figure 2.**
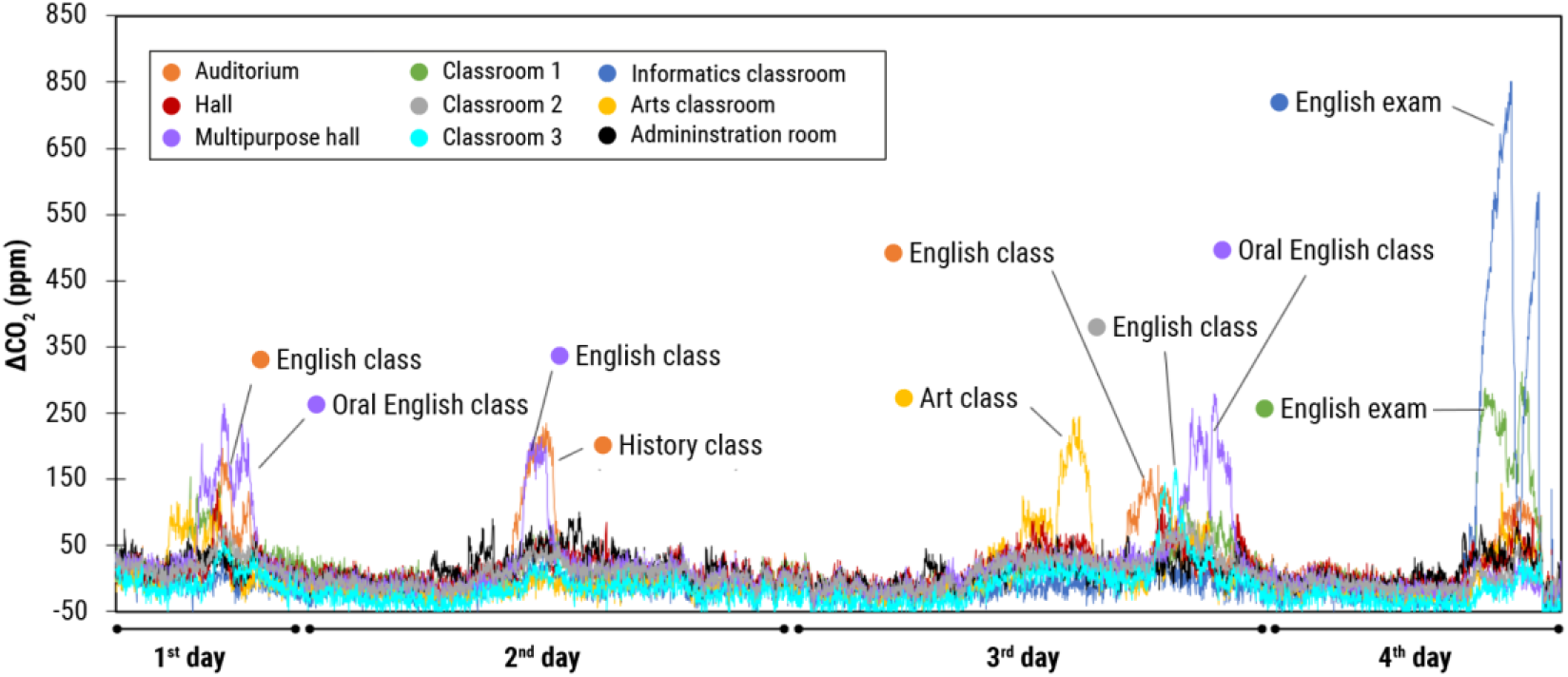
Increment of CO_2_ values recorded in the diagnostic period.

Comparing the ppm/person indicator in similar activities in different spaces makes it possible to determine the relative efficiency of air renewal in each space. For instance, in English classes (not oral), 9.2 ± 16.4, 13.1 ± 21.7, and 18.1 ± 21.3 ppm/person were achieved in Classroom 1, the Auditorium, and the Multipurpose Room, respectively. It indicates that air renewal is similar in all rooms, although slightly improved in Classroom 1. On the contrary, disparate ratios were obtained in the Cambridge exams in Classroom 1 and the Informatics Classroom. While in Classroom 1, the ratio was 12.2 ± 17.3 ppm/person, in the Informatics Classroom, a 46.9 ± 107.3 ppm/person was found. Again, this indicates more efficient air renewal inside the basement room.

Some gaps may go unnoticed during the diagnostic phase. Due to the atmospheric conditions or the activity during the diagnosis, the CO_2_ levels may be decreased. Although we recommend analyzing 3-4 critical days, subsequent days (e.g., 5-6 days) should be closely monitored to ensure representative results. For example, in the base case, on the monitoring days after diagnosis, higher CO_2_ concentrations were recorded in Classroom 1 than in other areas, even in similar conditions of capacity and activity.

### 3.2. Interpretation of CO_2_ measurements, methods to correct gaps, and monitoring

After detecting spaces that can be improved, it is necessary to implement efficient corrective measures that guarantee adequate air renewal. First, it is crucial to interpret the measurements and begin to rule out possibilities. In the case of Classroom 1, the following hypotheses were prepared:

- Hypothesis 1. The activities studied in the initial diagnosis were not representative, and the high activity in the classroom, so more CO_2_ is exhaled. Then, the following 12 English classes in Classroom 1, comprising the months of April to June, were analyzed. In the studied activities, no significant increase in CO_2_ was observed, except for the classes held in June, where an increase of 522 ppm was reached. Figure 3 shows the evolution of average CO_2_ from April to June, showing an increase in concentration in the latter month. However, CO_2_ exhalation in the records studied is similar, so this hypothesis is ruled out.
- Hypothesis 2. The weather conditions of the initial diagnosis were favorable for air renewal ratios. The weather influence on the ventilation ratios was evaluated by analyzing the incidences (values above 800 ppm for more than 30 minutes) recorded from April to July (regardless of the activity carried out). Of the 13 incidences found, 46 % (6) came from July, while 31 % (4) came from June, and the remaining 23% (3) from May. The climatic variation has affected the ventilation performance of the space. Working on days with less favorable weather will be necessary to test the corrective measures.
- Testing corrective measures. An experiment where the space was saturated with CO_2_ using dry ice was conducted, stabilized as a function of the amount evaporated. Two scenarios were evaluated with an activated HVAC system: with and without opening doors. Figure 4 shows the results obtained. With doors closed, it takes ∼1,600 ppm ∼37 minutes to reduce the CO_2_ concentration to ∼800 ppm, at 22 ppm/minute. While, with doors open, the reduction from ∼1,600 ppm to ∼800 ppm occurs in ∼17 minutes, at 47 ppm/minute. The opening of doors with the HVAC system active supposes a remarkable improvement of the ventilation in this classroom.

**Figure 3.**
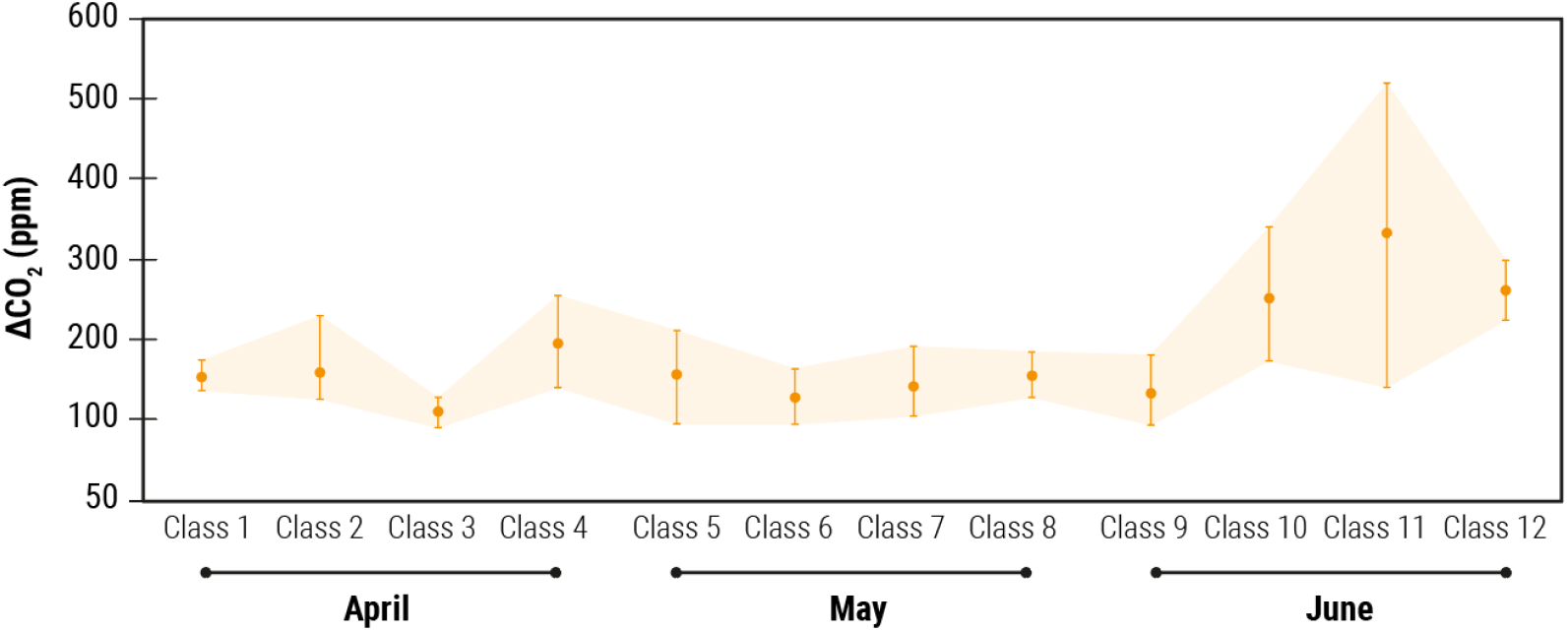
Evolution of average ΔCO_2_ in English classes from April to June.

**Figure 4.**
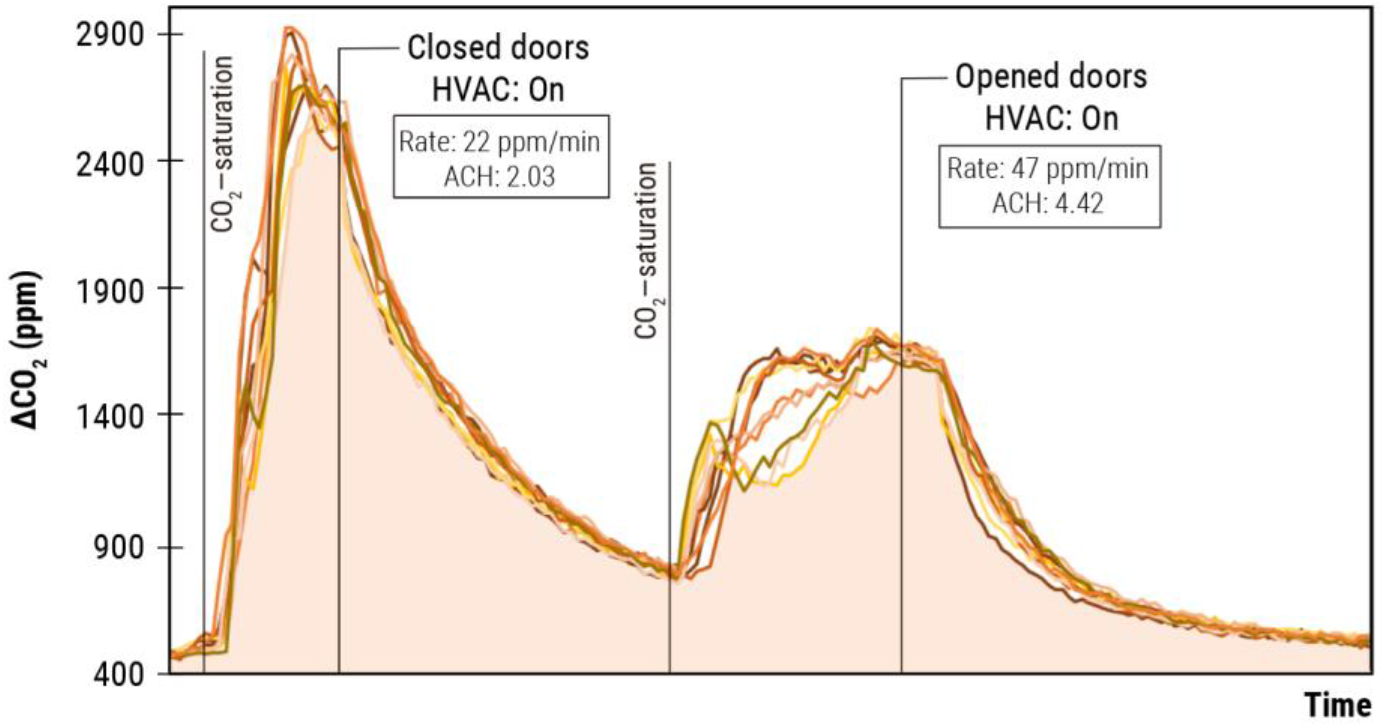
Results of the experiment with dry ice in Classroom 1, with and without open doors.

In addition, this method helps calculate actual air changes per hour (ACH) in space. The HVAC flow in Classroom 1 is 750 m^3^/s. For a volume of 363 m^3^ corresponding to the space, a theoretical ACH of 2.06 (door closed) is estimated. An ACH of 4.42 (with opened doors) and 2.03 (closed doors) has been determined by dry ice tests. It suggests that the tests have been carried out correctly and that the opening of the doors represents a considerable improvement in air renewal compared to the closed space.

Sometimes, the subsequent monitoring phase can be used to assess the benefit of the corrective actions imposed. In the base case, opening doors in Classroom 1 was enough to achieve adequate air renewal, even in the summer months. Ideally, during monitoring, it would be necessary to implement a system that alerts those responsible for the space if the established CO_2_ limits are exceeded. This strategy allows for preventing possible incidents and correcting CO_2_ levels exceeded in streaming, thus, guaranteeing adequate air quality.

### 3.3. Applying and validating the method in local businesses

To perform a broader application of the methodology to diagnose, correct, and monitor CO_2_ we performed a study in 40 retail stores. To this end we modified the phases to meet the pre-established requirements, and the absolute CO_2_ concentration was measured (instead of the outdoor-indoor CO_2_ increase). In addition, the ppm/person indicator could not be used since the businesses could not provide a continuous record of the occupancy. Initially, a preliminary characterization (diagnosis) of the participant businesses was conducted. 23 businesses with CO_2_ values above the maximum recommended (800 ppm) were identified. Eight of them did not require specific follow-up interventions as they were minimal and discrete increases. The other 15 participants were subjected to an exhaustive evaluation that served to design and implement a series of prevention measures that had to be implemented to improve air recirculation in the commercial spaces.

The implementation of corrective measures was modified from the original approach. Instead of *in-situ* experimenting, it was decided to assess the improvement of the proposed measures throughout the MCS stages. Thus, we evaluate the air renewal patterns from week to week. In our case, it was enough to agree on the terms with each person in charge through fortnight reports and phone calls. The presented strategy allowed us to reduce the costs of the project and to be able to evaluate several stores at the same time.

In the period studied (January-February 2022), the average CO_2_ of all businesses was 547 ± 87 ppm in the first fortnight of January, 559 ± 86 ppm in the second fortnight of January, 531 ± 82 ppm in the first fortnight of February, and 529 ± 89 in the second half of January (Figure 5a). Substantial differences were observed according to the type of store, as shown in Figure 5b. Specifically, the highest CO_2_ peaks were observed in Kids’ clothing stores (691 ± 209 ppm with maximums of 2,297 ppm). It may be associated with a limited opening of doors or windows to maintain the thermal comfort of the children. In contrast, florists reported the lowest CO_2_ levels (mean of 466 ± 47 ppm with a maximum of 874 ppm), possibly due to customers’ reduced time during service.

**Figure 5.**
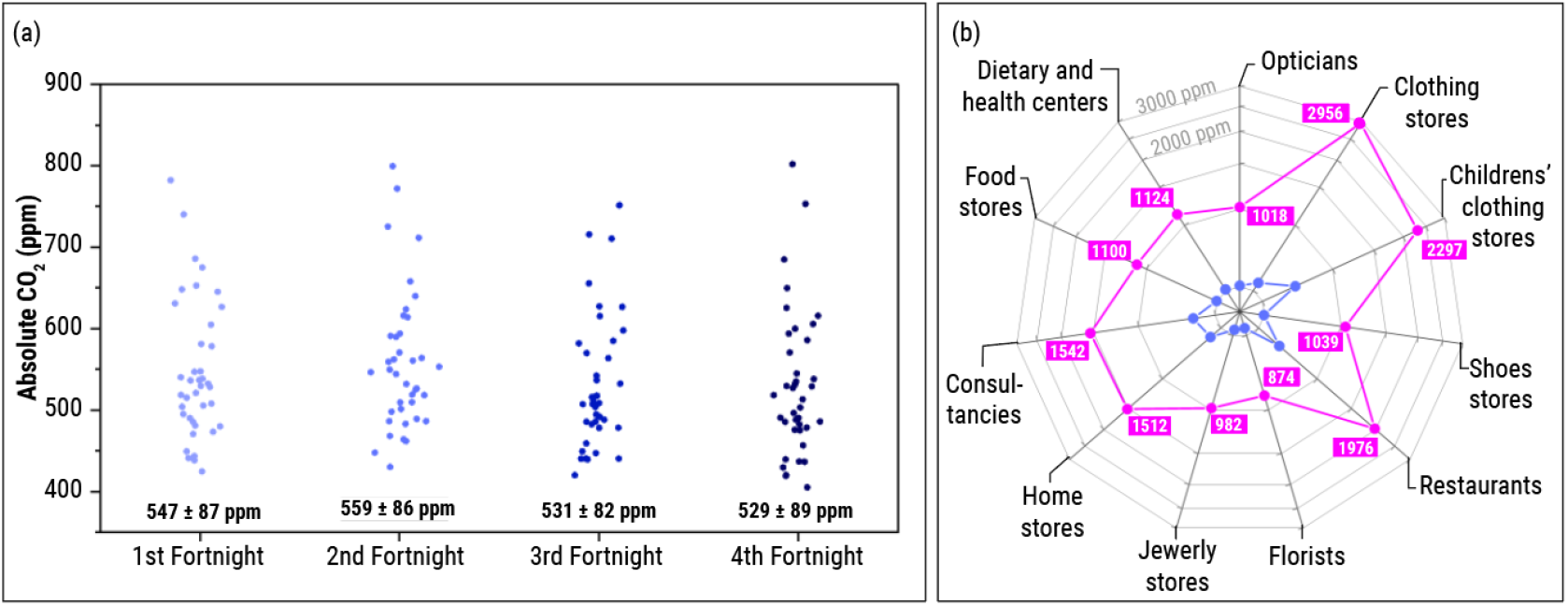
(a) Mean of CO_2_ value (inbox) of all businesses based on the fortnight and (b) mean of CO_2_ and maximum measurement depending on the type of business where the mean value (blue) and the maximum values (pink label) are shown.

A total of 15 businesses were selected for exhaustive monitoring. Among them, only five agreed to implement the corrective measures. The following sections describe the evolution of 3 establishments under the three typical scenarios:

1. Retail stores with good starting air quality. Initially, 17 shops showed a renewal of the air appropriate to the architecture of the space and the type of activity carried out. Three cases are shown below as an example. *Example 1)* In a shoe store, the limit of 800 ppm was not exceeded during the recorded period (Figure 6a). Specifically, it obtained an average CO_2_ value of 441 ± 42 ppm, reaching a maximum value of 776 ppm. *Example 2)* A food store was kept below 800 ppm (Figure 6b). Despite the large influx of customers, the levels maintained an average of 454 ± 39 ppm, reaching a maximum value of 782 ppm *Example 3)* In one of the clothing stores, the maximum established by the scientific community of 800 ppm was not exceeded (Figure 6c). On average, it reached a value of 469 ± 33 ppm, reaching a maximum value of 783 ppm.
2. Businesses with improvable air quality that did not implement corrective actions. Of the 23 stores that showed levels above 800 ppm during the first week, only 15 of them did so continuously and with values above 900 ppm. The remaining eight stores were recommended to maximize the frequency of opening doors and windows. This section shows the evolution of CO_**2**_ in a business that refused to establish the proposed corrective measures. *Example 1)* On average, a clothing store reached a value of 709 ± 295 ppm, reaching a maximum of 2956 ppm (Figure 6d). *Example 2)* A restaurant establishment reached an average value of 589 ± 139 ppm, reaching a maximum value of 1976 ppm (Figure 6e). *Example 3)* The mean of CO_**2**_ in a children’s clothing store was 762 ± 229 ppm, reaching a maximum of 2297 ppm (Figure 6f).
3. Spaces with improvable air quality that implemented corrective actions. After implementing the prevention measures (especially the reinforcement of natural ventilation), the reduction of CO_**2**_ in the establishments was substantial. Three cases are described below. The values of the first weeks concerning the final weeks are comparable, given that similar trends and averages are observed in the previous section. *Example 1)* In a clothing shop, the mean of CO_**2**_ was reduced from 605 ± 78 ppm (maximum of 1100 ppm) to 529 ± 78 ppm (745 ppm) after trying different measures. The increase in the frequency of door opening (Figure 6g) was more effective in the fourth fortnight. In this period, no values over 800 ppm were registered. *Example 2)* CO_2_ levels on average were reduced from 536 ± 89 ppm (maximum of 1001 ppm) to 485 ± 51 ppm (714 ppm) from the third fortnight in a food store, as shown in Figure 6h. *Example 3)* In a shoe store, the mean of CO_**2**_ was reduced from 528 ± 67 ppm (maximum of 946 ppm) to 473 ± 45 ppm (764 ppm). Although, at one point, 800 ppm was exceeded (Figure 6i).

**Figure 6.**
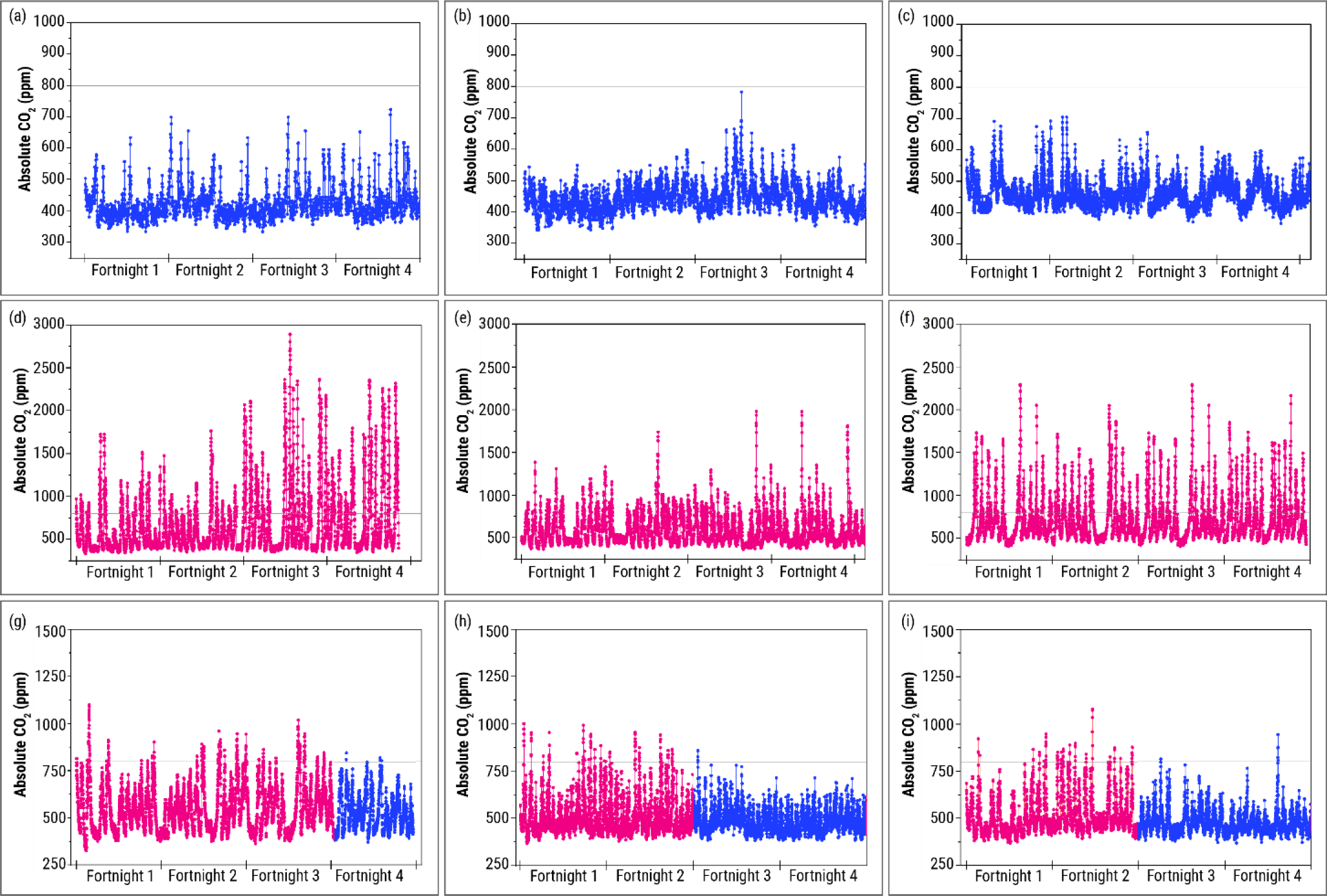
Evolution of CO_2_ levels in the monitoring period (January-February 2022) in (a-c) businesses with good starting air quality, (d-f) businesses with improvable air quality that did not implement corrective measures, and (g-i) businesses with improvable air quality that implemented corrective actions. The pink dots indicate the CO_2_ curves of stores with poor air renewal, while the blue dots represent CO2 curves of establishments with good air renewal.

## 4. Conclusions

This work presents and discusses a new methodology for measuring CO_2_ in shared spaces based on diagnosis, correction, and MCS (monitoring-control and surveillance phases). This methodology was validated in a subset of 40 local businesses in Zaragoza (Spain). The COVID-19 pandemic has pointed to the need to rethink the epidemiological control techniques. In particular, the increasing knowledge on airborne transmission of diseases makes it imperative to implement new public health strategies that guarantee that air quality follows specific standards. If the incorporation of these strategies proves effective, it will be possible to prevent numerous respiratory affections and potential new pandemics.

Briefly, the methodology can be sequenced in three stages:

1. Diagnosing. After training the responsible person, the diagnostic phase was carried out. A first assessment of the shared space is made by placing CO_2_ meters for a time interval long enough to address the maximum possible casuistry. Usually, 3 or 4 business days is enough. Ideally, placing more than 1 CO_2_ meter will let us know if the measurement represents the space. Additionally, it will bring preliminary -but no definitiveinformation about how the air renewal occurs in the shared space. After an in-depth analysis of the evolution of CO_2_ levels in the diagnosis phase, it is necessary to follow up in subsequent days (typically 6 or 7 days) to ensure that the diagnosis is representative of the space.
2. Correcting. Poor air renewal must be identified once an exhaustive analysis of the space has been carried out. Based on current scientific knowledge, a criterion of not exceeding 800 ppm for more than 20-30 minutes may be valuable. After detecting these incidences, it is necessary to look for the cause of the problem. It is helpful to raise possible hypotheses and try to rule them out to propose efficient corrective measures. Ideally, these proposed measures can be validated by conducting experiments (e.g., saturating with CO_2_ and analyzing the advantage of each one) or carrying out a detailed follow-up in the monitoring phase. Note that this phase can be performed linearly (e.g., after diagnosis), or it can also be implemented during monitoring if necessary.
3. MCS. A monitoring and follow-up phase is crucial to warn of any incident where the CO_2_ limit is exceeded. It is the only strategy that guarantees that good indoor air quality. It is possible that, in spaces where natural ventilation influences the space air renewal, the CO_2_ concentration fluctuates depending on the weather. In these cases, it may be necessary to re-implement the correction phase to maintain an adequate air renewal. Ideally, an alert system can be valuable for the monitored center, allowing it to prevent the accumulation of CO_2_ or modulate the capacity and, ultimately, correct the situation.

The results obtained from the validation in 40 retail stores in the center of Zaragoza (Spain) suggest that it is possible to implement measures that favor the renewal of shared air quickly. Opening doors and windows is one of the most straightforward and immediate methods applied in almost any situation. We have shown that the methodology is practical and can reduce the risk of transmission of respiratory diseases by aerosols by increasing air renovation. We hope this work contributes to laying the methodological foundations for CO_2_ measurement to become a valuable, standardized, and guaranteed tool.

## Data Availability

All data produced in the present work are contained in the manuscript

## Acknowledgments

We would like to express our gratitude to “Fundación Ibercaja” and “Federación de Empresarios de Comercio y Servicios de Zaragoza (ECOS Zaragoza)” for supporting this research and their invaluable collaboration. We thank Signos.io for help and support with the CO_2_ meters. We also would like to thank “Ayuntamiento de Zaragoza” and “Instituto de Investigación Sanitaria Aragón” for the received support.

